# Pre- and asymptomatic viral shedding in high-risk contacts of monkeypox cases: a prospective cohort study

**DOI:** 10.1101/2022.11.23.22282505

**Authors:** Isabel Brosius, Christophe Van Dijck, Jasmine Coppens, Leen Vandenhove, Eugene Bangwen, Fien Vanroye, Jacob Verschueren, Sabine Zange, Joachim Bugert, Johan Michiels, Emmanuel Bottieau, Patrick Soentjens, Johan van Griensven, Chris Kenyon, Kevin K. Ariën, Marjan Van Esbroeck, Koen Vercauteren, Laurens Liesenborghs, the ITM MPX consortium

## Abstract

Epidemiological data suggest that clade IIb monkeypox virus (MPXV) is readily transmitted during sexual contact, even before symptom onset. However, presymptomatic shedding of MPXV remains to be demonstrated. Here, we prospectively followed up 25 individuals after high-risk exposure to MPXV. Daily anorectal, genital, and saliva samples and weekly blood and oropharyngeal samples were collected along with clinical information. During follow-up, 12/18 (66.0%) sexual and 1/7 (14.0%) non-sexual contacts showed evidence of MPXV infection by PCR, five of whom had low viral loads and no typical MPXV symptoms. In 5/6 (83.3%) patients with typical symptoms, viral DNA was detected as early as four days before symptom onset and in three of them, replication-competent virus was found. These findings emphasize the high risk of MPXV transmission during sexual contact and confirm the existence of presymptomatic viral shedding of MPXV. Sexual contacts of an MPXV-infected partner should abstain from sex irrespective of symptoms.

## Introduction

Since May 2022, an outbreak of monkeypox (MPX) has caused more than 80,000 laboratory-confirmed cases across the world, predominantly among men who have sex with men (MSM).^1^ This current epidemic is caused by variant B.1 of the subclade IIb of MPX virus (MPXV), and, in contrast to previous outbreaks, is driven uniquely by human-to-human transmission, especially through sexual contact.^2^ In addition, the clinical presentation in this global outbreak differs from what was commonly reported before 2022. Lesions often predominate at the site of inoculation and frequently involve mucosal membranes, resulting in proctitis, urogenital symptoms, or tonsillitis.^2^

Based on data from previous outbreaks, most guidelines, including those issued by WHO, ECDC, and CDC, consider patients infectious from the start of symptoms until the complete healing of skin lesions.^3,4^ For that reason, public health messaging has mainly focused on awareness of symptoms, early diagnosis and isolation of symptomatic cases.

However, it was recently demonstrated that asymptomatic MPX infections could play an important role in transmission.^5,6^ Furthermore, recent epidemiological data suggest that presymptomatic transmission also occurs and could be responsible for about half of all infections.^7^ Presymptomatic transmission can take place when viral shedding precedes clinical symptoms and often has a major impact on epidemic dynamics, as observed during the COVID-19 outbreak.^8^ Nevertheless, presymptomatic shedding of MPXV, the body sites from which it may occur and its timing with relation to symptom onset remain elusive.

To study the risk of infection after exposure to MPXV and the natural history of the early phase of MPXV infection, we performed a detailed follow-up of high-risk contacts of clade IIb MPXV patients. Here, we described their clinical and virological characteristics from exposure until diagnosis.

## Results

Between June 24 and July 31, 2022, we prospectively followed up 25 high-risk contacts of confirmed MPX cases. All participants except one (96.0%) self-identified as MSM; the median age was 43 years (IQR 36 – 51, **Table 1**). Eighteen (72.0%) participants reported having had sexual contact with an index case. Seven (28.0%) were non-sexual high-risk contacts: five were household contacts and two had prolonged skin-to-skin contact with an MPX-confirmed case. The median time between last exposure and inclusion was 8.5 (IQR 5 – 10) days. Five participants received post-exposure vaccination (PEV) and six reported being vaccinated against smallpox during childhood.

**Table 1.**
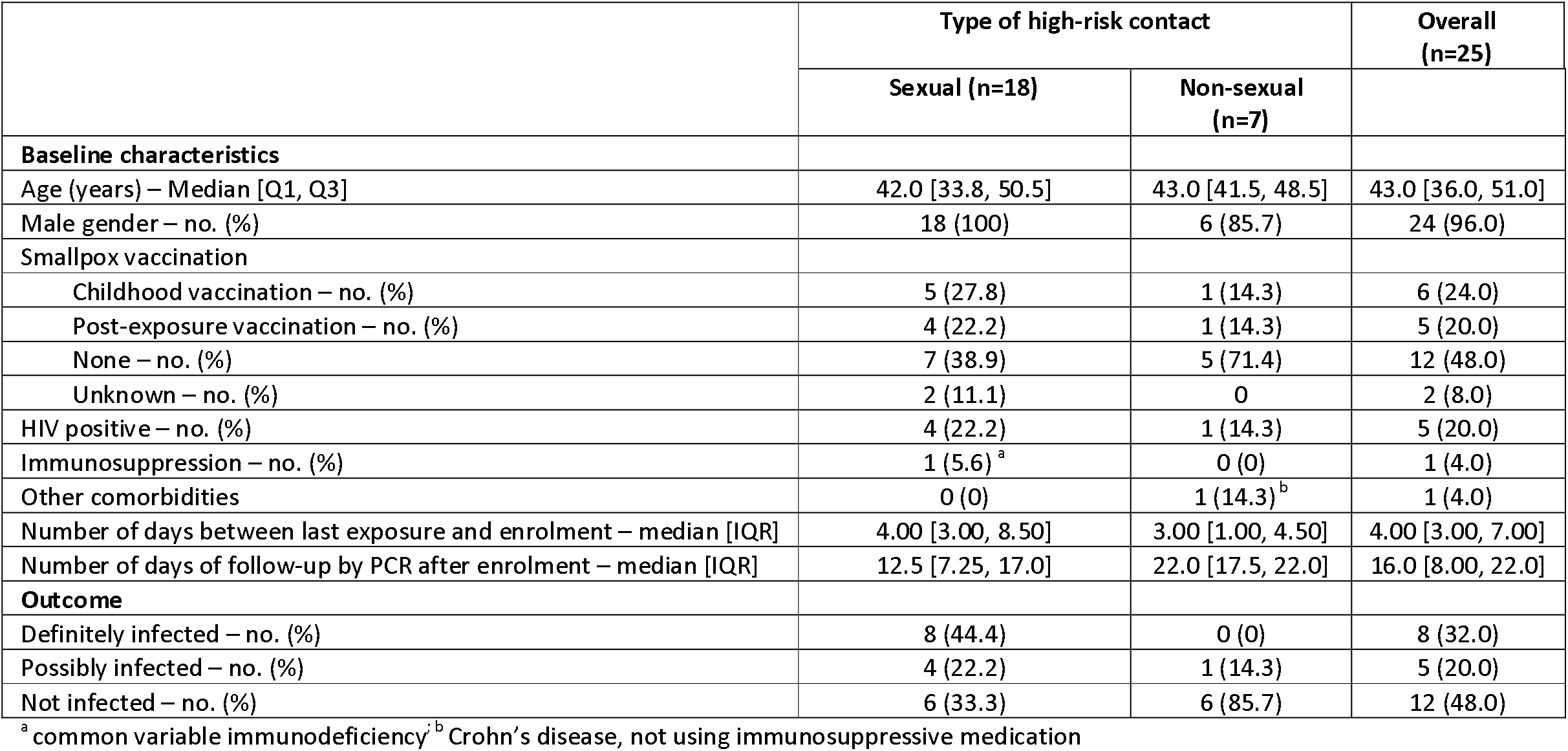
Participant characteristics and clinical outcome by type of high-risk contact.

|  | Type of high-risk contact |  | Overall<br>(n=25) |
| --- | --- | --- | --- |
|  | Sexual (n=18) | Non-sexual<br>(n=7) |  |
| Baseline characteristics |  |  |  |
| Age (years) – Median [Q1, Q3] | 42.0 [33.8, 50.5] | 43.0 [41.5, 48.5] | 43.0 [36.0, 51.0] |
| Male gender – no. (%) | 18 (100) | 6 (85.7) | 24 (96.0) |
| Smallpox vaccination |  |  |  |
| Childhood vaccination – no. (%) | 5 (27.8) | 1 (14.3) | 6 (24.0) |
| Post-exposure vaccination – no. (%) | 4 (22.2) | 1 (14.3) | 5 (20.0) |
| None – no. (%) | 7 (38.9) | 5 (71.4) | 12 (48.0) |
| Unknown – no. (%) | 2 (11.1) | 0 | 2 (8.0) |
| HIV positive – no. (%) | 4 (22.2) | 1 (14.3) | 5 (20.0) |
| Immunosuppression – no. (%) | 1 (5.6)<sup>a</sup> | 0 (0) | 1 (4.0) |
| Other comorbidities | 0 (0) | 1 (14.3)<sup>b</sup> | 1 (4.0) |
| Number of days between last exposure and enrolment – median [IQR] | 4.00 [3.00, 8.50] | 3.00 [1.00, 4.50] | 4.00 [3.00, 7.00] |
| Number of days of follow-up by PCR after enrolment – median [IQR] | 12.5 [7.25, 17.0] | 22.0 [17.5, 22.0] | 16.0 [8.00, 22.0] |
| Outcome |  |  |  |
| Definitely infected – no. (%) | 8 (44.4) | 0 (0) | 8 (32.0) |
| Possibly infected – no. (%) | 4 (22.2) | 1 (14.3) | 5 (20.0) |
| Not infected – no. (%) | 6 (33.3) | 6 (85.7) | 12 (48.0) |

<sup>a</sup> common variable immunodeficiency; <sup>b</sup> Crohn's disease, not using immunosuppressive medication

Participants performed daily self-sampling of anorectal swabs, genital swabs, and saliva and were asked to keep a symptom diary (**Fig. 1A**). In addition, participants attended weekly visits for clinical examination and collection of blood and oropharyngeal swabs. Those who developed typical MPX symptoms (skin lesions, proctitis, urethritis, tonsillitis) were evaluated ad hoc at the clinic. Follow-up ended if a participant had typical MPX symptoms and a positive MPXV-PCR (Cycle threshold - Ct-value < 34). Overall, participants were followed up for a median of 16 (IQR 8 – 22) days, i.e. until day 23 (IQR 14 – 26) after their last high-risk contact.

**Fig. 1.**
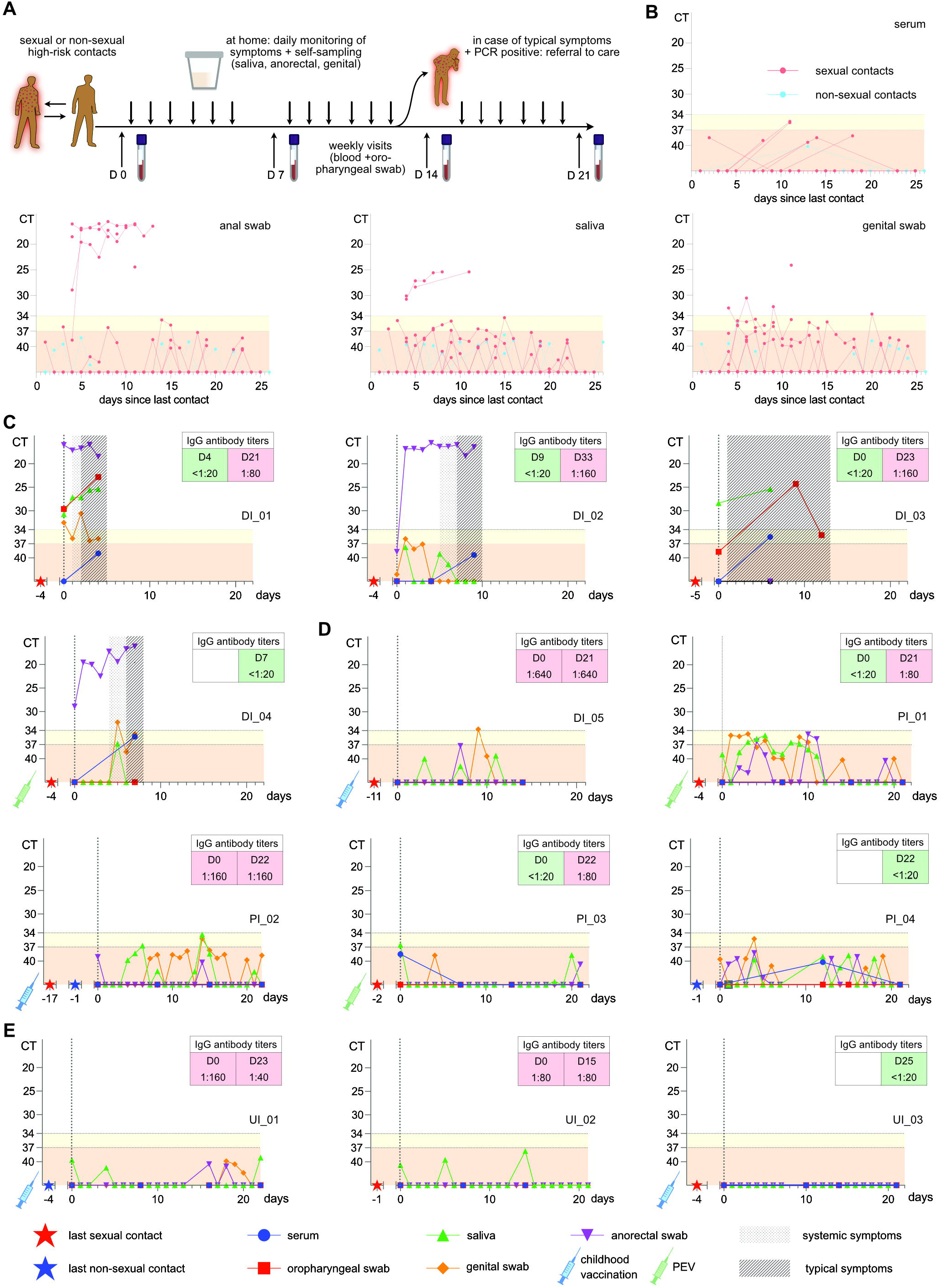
**(A)** Schematic overview of the study design. **(B)** Overview of MPXV-PCR results of all sexual and non-sexual contacts, in relation to the day of last exposure (day 0). Dots indicate PCR cycle threshold (Ct) values of individual samples; lines indicate individual participants. Ct-values < 34 (white area), 34-37 (yellow area) and ≥ 37 (red area) are considered positive, weakly positive and negative, respectively. **(C-E)** Serology and MPXV-PCR results in a selection of the most illustrative cases; **(C)** selected symptomatic cases with presymptomatic shedding, **(D)** selected asymptomatic or atypical cases, **(E)** selected uninfected participants. Symbols indicate the day of last sexual or non-sexual exposure and vaccination status, shaded areas indicate the presence of systemic and typical monkeypox symptoms. Individual participant identification consists of category: DI = definitely infected, PI = possibly infected, or UI = uninfected and participant number. D denotes day.

A total of 1,108 samples were collected and analyzed by MPXV-PCR (**Supplementary Table 1**), including 323 saliva, 323 anorectal, 312 genital, 70 oropharyngeal, 66 serum, and 14 skin samples. A total of 184 (16.6%) samples were MPXV-PCR positive, of which 142 (77.1%) showed Ct-values of 34 and higher, often in the absence of symptoms. Considering the recent reports of false positive MPXV-PCR results in patients with a low clinical suspicion of MPX,^9^ we defined two Ct-value cutoffs: one highly specific (Ct 34) and another highly sensitive (Ct 37). The latter cutoff was based on a specificity analysis of saliva samples from 52 MPXV-uninfected controls. In addition, samples with Ct-values between 34 and 37 underwent confirmation testing by repeating the MPXV-PCR and a PCR melting curve analysis. Unconfirmed results were reported as negative. Overall, anorectal samples more often had a Ct-value < 34 (7.7% of samples), compared to saliva (2.5%) and genital swabs (1.3%). In contrast, saliva and genital swabs were more often weakly positive (3.1% and 4.1% of samples, respectively) compared to anorectal swabs (1.2%) (**Fig 1A**, Supplementary Fig. 1).

Based on the two MPXV-PCR Ct-value cutoffs, the infection status of individual participants was defined as one of three outcomes: definitely infected, possibly infected, or uninfected (**Box 1**). Using these criteria, we found that a high proportion (n=12/18, 66.7%) of sexual contacts were definitely (n=8) or possibly infected (n=4, **Table 1**). In contrast, among the non-sexual contacts, only one out of seven (14.2%) was possibly infected, and none were definitely infected (*p*= .03 for comparing infection status between sexual and non-sexual contacts, Fisher’s Exact Test). Orthopoxvirus serology confirmed the patients’ outcome in 12/25 (48.0%) cases, was uninterpretable in 10/25 (40.0%) cases (either due to PEV or positive serology at baseline) and contradicted PCR results in 3/25 (12.0%) cases (two definitely infected and one possibly infected case without documented seroconversion). (**Supplementary Table 1**).

### BOX 1

**Infection status outcome definitions**

- definitely infected=at least one sample with a MPXV-PCR Ct-value < 34
- possibly infected= at least one sample with a MPXV-PCR Ct-value ≥ 34 to < 37
- uninfected= all MPXV-PCR Ct-values ≥ 37 Ct=cycle threshold; MPXV=monkeypox virus

Among the eight definitely infected cases, six (75.0%) developed typical MPX symptoms, one (12.5%) had only fever and another (12.5%) only fatigue (**Table 2**). Typical symptoms included skin rash (n=4), proctitis (n=2), and tonsillitis (n=2), and were preceded by a prodromal phase in all cases. **Fig. 1C-E** depicts the evolution of symptoms and PCR results for selected illustrative cases. Remaining cases are presented in **Supplementary Fig. 2**. In 5/6 (83.3%) typical cases, viral DNA was detected one (n=3) and four (n=2) days before the onset of any symptoms in oropharyngeal (n=1), saliva (n=2), anorectal (n=4), and genital (n=3) samples. In 3/4 anorectal samples, we were able to culture the virus during the presymptomatic phase. The fourth anorectal sample had insufficient volume for culture.

**Table 2:** Clinical presentation and viral DNA detection by outcome.

|  | Definitely<br>infected (n =<br>8) | Possibly infected<br>(n = 5) | Uninfected (n =<br>12) | Overall (n =<br>25) |
| --- | --- | --- | --- | --- |
| Asymptomatic – no. (%) | 0 (0) | 2 (40.0) | 7 (58.3) | 9 (36.6) |
| Fever or night sweats only – no. (%) | 1 (12.5) | 1 (20.0) | 0 (0) | 2 (8.0) |
| Other<sup>a</sup> symptoms only – no. (%) | 1 (12.5) | 2 (40.0) | 4 (33.3) | 7 (28.0) |
| Typical<sup>b</sup> symptoms – no. (%) | 6 (75.0) | 0 (0) | 1 (8.3)<sup>c</sup> | 7 (28.0) |
| Skin rash – no. (%) | 4/6 (66.7) | NA | 0/1 (0) | 4/7 (57.1) |
| Proctitis – no. (%) | 2/6 (33.3) | NA | 0/1 (0) | 2/7 (28.6) |
| Urethritis – no. (%) | 0/6 (0) | NA | 0/1 (0) | 0/7 (0) |
| Tonsillitis – no. (%) | 2/6 (33.3) | NA | 1/1 (100)<sup>c</sup> | 3/7 (42.9) |
| Preceded by fever or other<sup>a</sup> symptoms – no. (%) | 6/6 (100) | NA | 0/1 (0) | 6/7 (85.7) |
| Preceded by pre-symptomatic MPXV DNA detection<br>(Ct-value < 37) – no. (%) | 5/6 (83.3) | NA | NA | 5/6 (83.3) |
| Number of days between most recent contact and first<br>positive MPXV DNA detection (Ct-value < 37) – median<br>(IQR) | 5.00 [4.00,<br>9.50] | 5.0 [5.0, 12.0] | NA | 5.0 [4.0, 11.0] |
| Number of days between most recent contact and first<br>symptoms – median (IQR) | 8.0 [4.5, 10.0] | 10.0 [6.5, 13.5] | 9.00 [6.00, 9.00] | 8.5 [5.0, 10.0] |

MPXV = monkeypox virus; NA = not applicable, IQR = interquartile range, Ct = PCR cycle threshold
<sup>a</sup> other symptoms: headache, fatigue, mild sore throat without tonsillitis on clinical exam
<sup>b</sup> typical symptoms: classical monkeypox skin lesions, proctitis, urethritis, tonsillitis
<sup>c</sup> case UI\_06 was diagnosed with a respiratory infection including tonsillitis due to COVID-19 during follow-up

Of the five possibly infected cases, none developed typical MPX symptoms. Two were asymptomatic, one had only night sweats, and two had other symptoms (headache, n=1; sore throat without tonsillitis, n=2).

Overall, patients without typical symptoms (two definitely infected, five possibly infected) had significantly lower viral loads compared to participants with typical symptoms (median of lowest recorded Ct-value 17.1 *vs* 34.8, *p*=.003, Mann Whitney test). Infected patients without typical symptoms tended to be more often vaccinated against smallpox, either during childhood (n = 2/7) or by PEV (n=3/7) compared to participants with typical symptoms (1/6 received PEV), although the difference was not statistically significant (*p*=.103, Fisher’s Exact Test).

## Discussion

Our extensive follow-up of high-risk contacts provides unique new insights into the early stages of MPX disease. First, our findings indicate that the risk of infection after exposure to clade IIb MPXV through sexual contact is much higher than previously appreciated. In contrast, the risk for household and other non-sexual contacts is low. Unfortunately, due to the waning epidemic in Belgium after July 2022, we could not recruit the predefined sample size of 140 participants, which would allow us to estimate the risk of infection more accurately.

Second, our data demonstrate that even though skin lesions and proctitis were common features in MPX cases reported during the 2022 global outbreak,^2^ such clinical presentations may be less common than generally assumed, as less than half of the infected cases in our study presented with typical monkeypox symptoms, and only one third had skin lesions. Notably, the atypical cases reported here generally had low viral loads, and most were vaccinated either through PEV or during childhood. They might, therefore, have been able to suppress viral replication and the development of full-blown disease. However, it is noteworthy that we faced similar difficulties as others when interpreting weakly positive MPXV-PCR results.^9^ Confirming the participants’ infection status by serology proved problematic because many cases had or developed orthopoxvirus IgG after childhood or post-exposure smallpox vaccination, and others remained seronegative despite MPXV-PCR Ct-values < 37. Overall, the exact clinical and epidemiological significance of cases with low levels of detectable viral DNA has yet to be determined.

Last, we detected presymptomatic MPXV DNA and even replication-competent virus in five out of six participants with typical MPX symptoms, as early as 4 days before symptom onset. In reality, presymptomatic shedding might start even earlier, as seven cases in our study were already PCR-positive at inclusion. The existence of presymptomatic transmission was suggested by an epidemiological study of surveillance and contact tracing data in the United Kingdom, which found that the median serial interval in 79 case-contact pairs was shorter than the median incubation period of 54 cases in the data set and that exposure of the contact took place during the presymptomatic phase of the index case in 10 out of 13 case-contact pairs.^7^ We now provide biological evidence that infected individuals are infectious during the presymptomatic phase, corroborating the epidemiological evidence of presymptomatic transmission. Moreover, we show that anorectal and, to a lesser extent, saliva and genital self-sampling are useful to detect such early-stage infections.

In conclusion, our data emphasize the high risk of infection during sexual contact, even in the presymptomatic phase. Contacts of an MPX cases should be aware of the possibility of presymptomatic viral transmission and advised to abstain from sex irrespective of symptoms.

## Online methods

### Study design and participants

In this study, we prospectively followed up individuals who had high-risk contact with a confirmed monkeypox (MPX) patient. Participants were recruited in two ways: either by referral through their index cases, or when they presented for post-exposure vaccination. High-risk contact was defined as either sexual contact, prolonged (> 15 minutes) skin-to-skin contact with an MPX patient with active skin lesions, or living in the same household as an MPX patient. Adult individuals were included in this study if their contact occurred in the 21 days prior to recruitment and if they provided written informed consent for study participation.

Study participants attended a predefined schedule of clinic visits, including one baseline visit and weekly follow-up visits. At baseline, we recorded medical history including smallpox vaccination status, date and type of contact with the index case. At every visit, symptoms were recorded through a standardized questionnaire, clinical signs of MPX were assessed by a thorough physical examination, and the following samples were collected: blood, saliva, oropharyngeal swabs, genital swabs, anorectal swabs, and swabs from skin lesions if applicable. Between study visits, participants completed a daily questionnaire on symptoms, and self-collected saliva, genital and anorectal samples at home. Follow-up was ceased maximum 21 +/−2 days after inclusion or as soon as any sample was MPXV-PCR positive with Ct-value <34 in a participant with typical MPX symptoms.

A sample size of 140 participants was predetermined to estimate the infection risk with a 95% confidence interval and a 5% margin of error. The hypothesized infection risk among contacts was hypothesized to be 10%. However, due to the waning of MPX epidemic after July, this sample size was not reached.

The study was conducted in accordance with the International Conference on Harmonization Guidelines for Good Clinical Practice and the Declaration of Helsinki and registered on ClinicalTrials.gov (NCT05443867). The protocol was approved by the Institutional Review Board of the Institute of Tropical Medicine (ITM) (1604/22) and by the Ethics Committee of the Antwerp University Hospital (2022-3571). The full study protocol is provided in the Supplementary Material section. Anonymized raw data will be made available upon request according to ITM’s data sharing policy.

### Sampling and sample handling

Blood (BD Vacutainer®, BD Benelux NV, Erembodegem), saliva (15 mL Safe-Lock Tubes, Eppendorf Belgium NV-SA, Aarschot) and all study swabs (Eswab, Copan Diagnostics, Brescia) were collected during clinic visits by a trained physician or nurse and were processed immediately. Home-based samples were collected using the same type of swabs and tubes, which were prelabelled for this purpose. Home-based samples were packaged in appropriate packaging material for storage in the participant’s refrigerator and were brought to the clinic at the next study visit.

### Laboratory procedures

The MPXV-PCR used in this study was an in-house PCR targeting the MPXV-TNF receptor gene carried out on the Applied Biosystems QuantStudio PCR system, as described previously.^5,10^ To confirm samples with MPXV-PCR Ct-values between 34 and 37, MPXV-PCR testing was repeated and melting curve assays were performed with the same MPXV primer set. Viral culture was performed as described previously on a subset of MPXV-PCR positive samples with Ct-value < 30.^5^ Orthopoxvirus serology was performed at the end of follow-up for all participants and at baseline for participants with positive end-of-follow-up serology (IgG titer ≥ 1:20). An in-house assay detecting anti-orthopoxvirus IgG at the Bundeswehr Institute of Microbiology was used.^11^

To assess the specificity of the MPXV-PCR and define Ct cut-off values, we collected saliva samples from 52 healthy volunteers who provided written informed consent. Samples were analyzed in triplicate and any positive PCR was considered a false positive result. Nineteen out of 156 (12.2%) analyses were positive, with a median Ct-value of 39.5 (range 39.3 to 42.6) (**Supplementary Table 1** and Supplementary Fig. 1.). A cutoff Ct-value of 37 was chosen to provide an additional margin of 2 Ct-values to guarantee adequate specificity. A second Ct-value cutoff with higher specificity was chosen at 34, based on recent reports of false positive MPXV-PCR results in patients with a low clinical suspicion of MPX.^9^

## Statistical analyses

Baseline characteristics and outcome variables were described as counts and proportions for categorical variables and means or medians with interquartile range for continuous variables. A two-sided Fisher’s exact test was used to compare proportions and a two-sided Mann Whitney U test to compare continuous variables. A p-value ≤ .05 was considered significant. Analyses were done with SPSS version 28.0.1.1 (IBM SPSS Statistics) and Prism version 9.4.1 (GraphPad Software, LLC). Figure panels and artwork were created with Affinity Designer version 2.0.0 (Serif Software).

## Data Availability

The data supporting the findings of this publication are retained at the Institute of Tropical Medicine (ITM), Antwerp and will not be made openly accessible due to ethical and privacy concerns. According to the ITM research data sharing policy, only fully anonymised data can be shared publicly. The data are de-identified (using a unique patient code) but not fully anonymised and it is not possible to fully anonymise them due to the longitudinal nature of the data. Data can however be made available after approval of a motivated and written request to the ITM at. The ITM data access committee will verify if the dataset is suitable for obtaining the study objective and assure that confidentiality and ethical requirements are in place.

## Acknowledgments

This work was funded by the Agency of Care and Health and the Department of Economy, Science, and Innovation of the Flemish government. IB and EB are members of the Institute of Tropical Medicine’s Outbreak Research Team, which is as also financially supported by the Department of Economy, Science, and Innovation

## Author contributions

I.B., L.L., M.V.E., K.V., J.v.G., P.S., E.Bo. and C.K. conceptualized the study. I.B., L.L. and L.V. managed the participants clinically. M.V.E., K.V., F.V., J.V., K.K.A., E.B. and J.C. supervised and coordinated the laboratory analyses at the Institute of Tropical Medicine. S.Z. and J.J.B. supervised and coordinated the laboratory analyses at the Bundeswehr Institute of Microbiology. J.C. and E.Ba. performed the testing at the Institute of Tropical Medicine. I.B., L.L., J.C. and C.V.D. analyzed the data. L.L., P.S. and J.v.G. secured funding. I.B., C.V.D. and L.L. wrote the first draft of the manuscript, with revision by K.V. and M.V.E. I.B. and C.V.D. contributed equally.

All authors reviewed and approved the final version of the manuscript.

## Online supporting material

### Supplementary table

**Supplementary Table 1:**
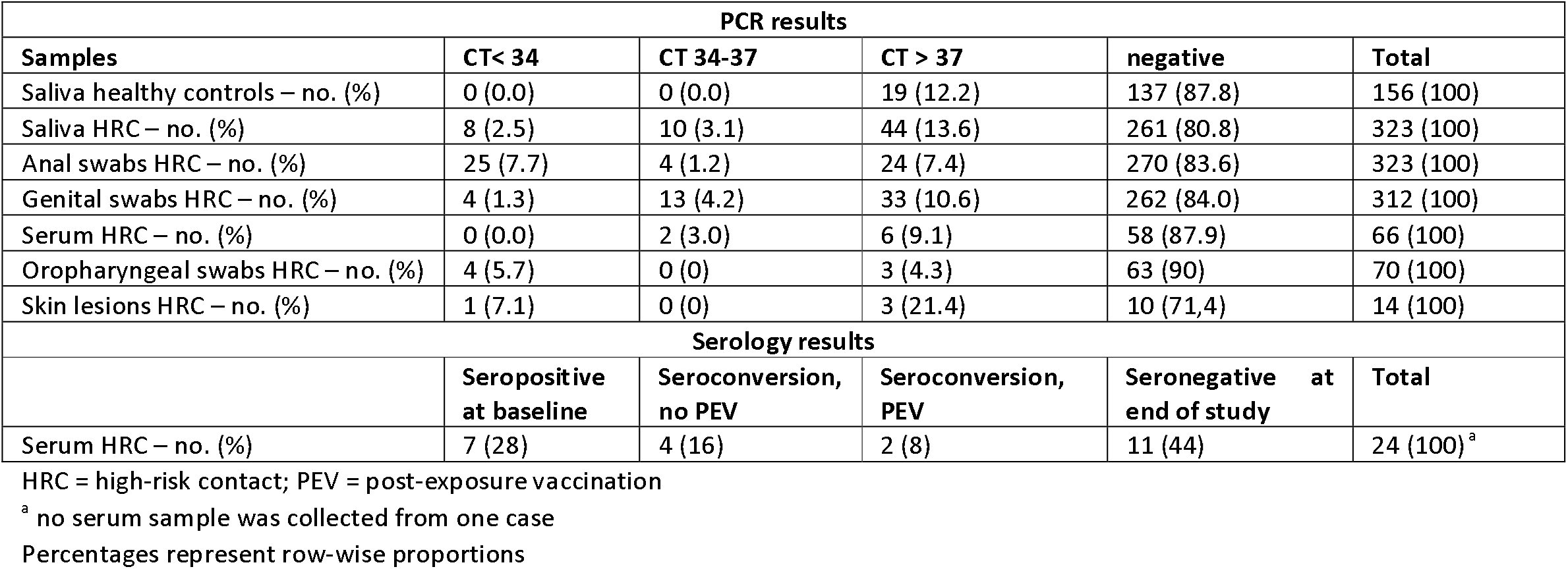
Samples and results of PCR and serology.

| PCR results |  |  |  |  |  |
| --- | --- | --- | --- | --- | --- |
| Samples | CT< 34 | CT 34-37 | CT > 37 | negative | Total |
| Saliva healthy controls – no. (%) | 0 (0.0) | 0 (0.0) | 19 (12.2) | 137 (87.8) | 156 (100) |
| Saliva HRC – no. (%) | 8 (2.5) | 10 (3.1) | 44 (13.6) | 261 (80.8) | 323 (100) |
| Anal swabs HRC – no. (%) | 25 (7.7) | 4 (1.2) | 24 (7.4) | 270 (83.6) | 323 (100) |
| Genital swabs HRC – no. (%) | 4 (1.3) | 13 (4.2) | 33 (10.6) | 262 (84.0) | 312 (100) |
| Serum HRC – no. (%) | 0 (0.0) | 2 (3.0) | 6 (9.1) | 58 (87.9) | 66 (100) |
| Oropharyngeal swabs HRC – no. (%) | 4 (5.7) | 0 (0) | 3 (4.3) | 63 (90) | 70 (100) |
| Skin lesions HRC – no. (%) | 1 (7.1) | 0 (0) | 3 (21.4) | 10 (71.4) | 14 (100) |
| Serology results |  |  |  |  |  |
|  | Seropositive at baseline | Seroconversion, no PEV | Seroconversion, PEV | Seronegative at end of study | Total |
| Serum HRC – no. (%) | 7 (28) | 4 (16) | 2 (8) | 11 (44) | 24 (100)<sup>a</sup> |

HRC = high-risk contact; PEV = post-exposure vaccination
<sup>a</sup> no serum sample was collected from one case
Percentages represent row-wise proportions

### Supplementary Figure Legend

**Supplementary Figure 1:** Results of MPXV-PCR by sample type. Ct value = MPXV-PCR cycle threshold value

**Supplementary Figure 2:** Serology and MPXV-PCR results in the remaining cases, not represented in Fig. 1 (A) definitely infected, symptomatic cases; (B) possibly infected, symptomatic case; (C) uninfected cases. Symbols indicate the day of last sexual or non-sexual exposure and vaccination status, shaded areas indicate the presence of systemic and typical monkeypox symptoms. Participant identification consists of category DI = definitely infected, PI = possibly infected, or UI = uninfected and participant number. D denotes day.

## ITM MPX consortium

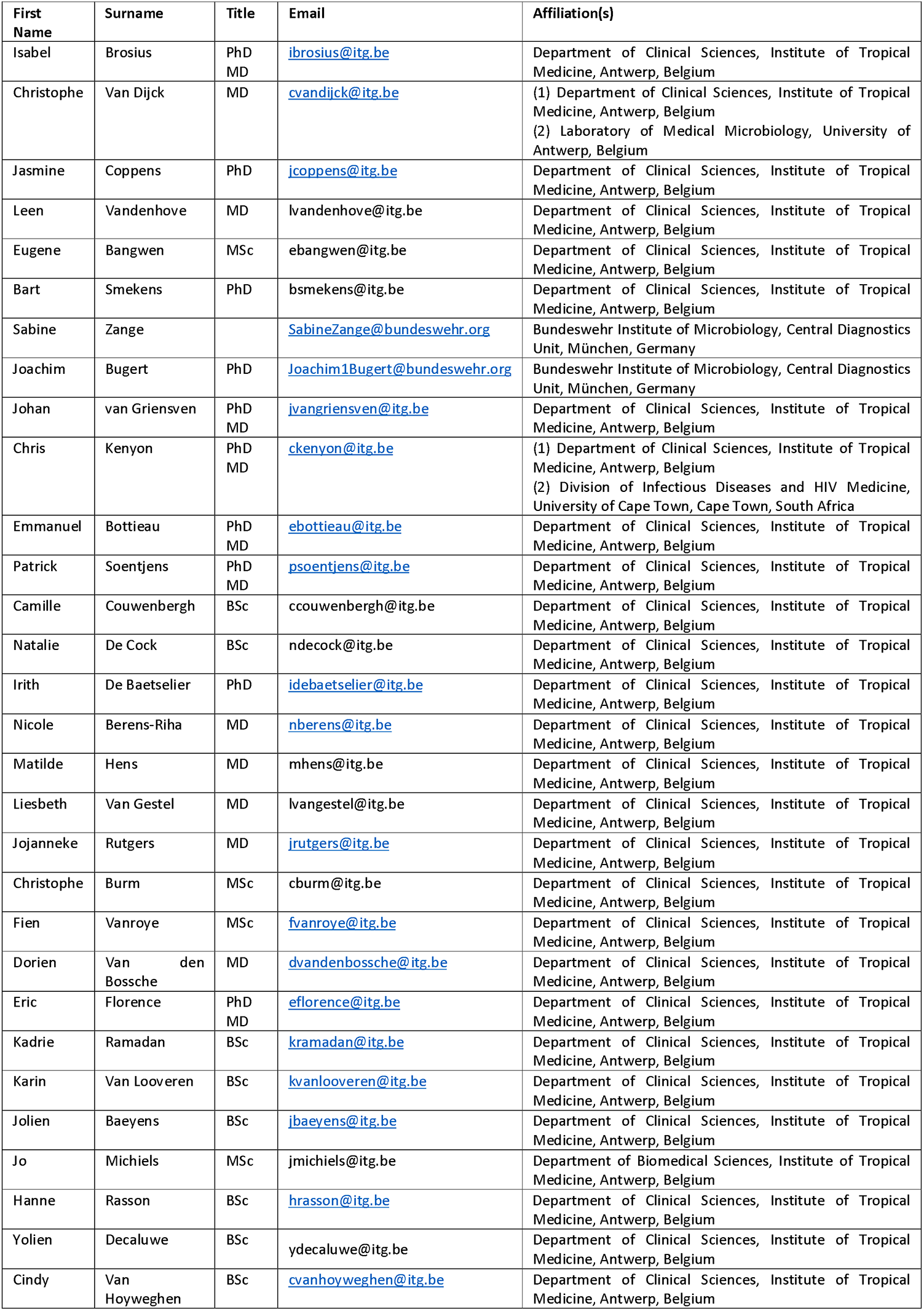
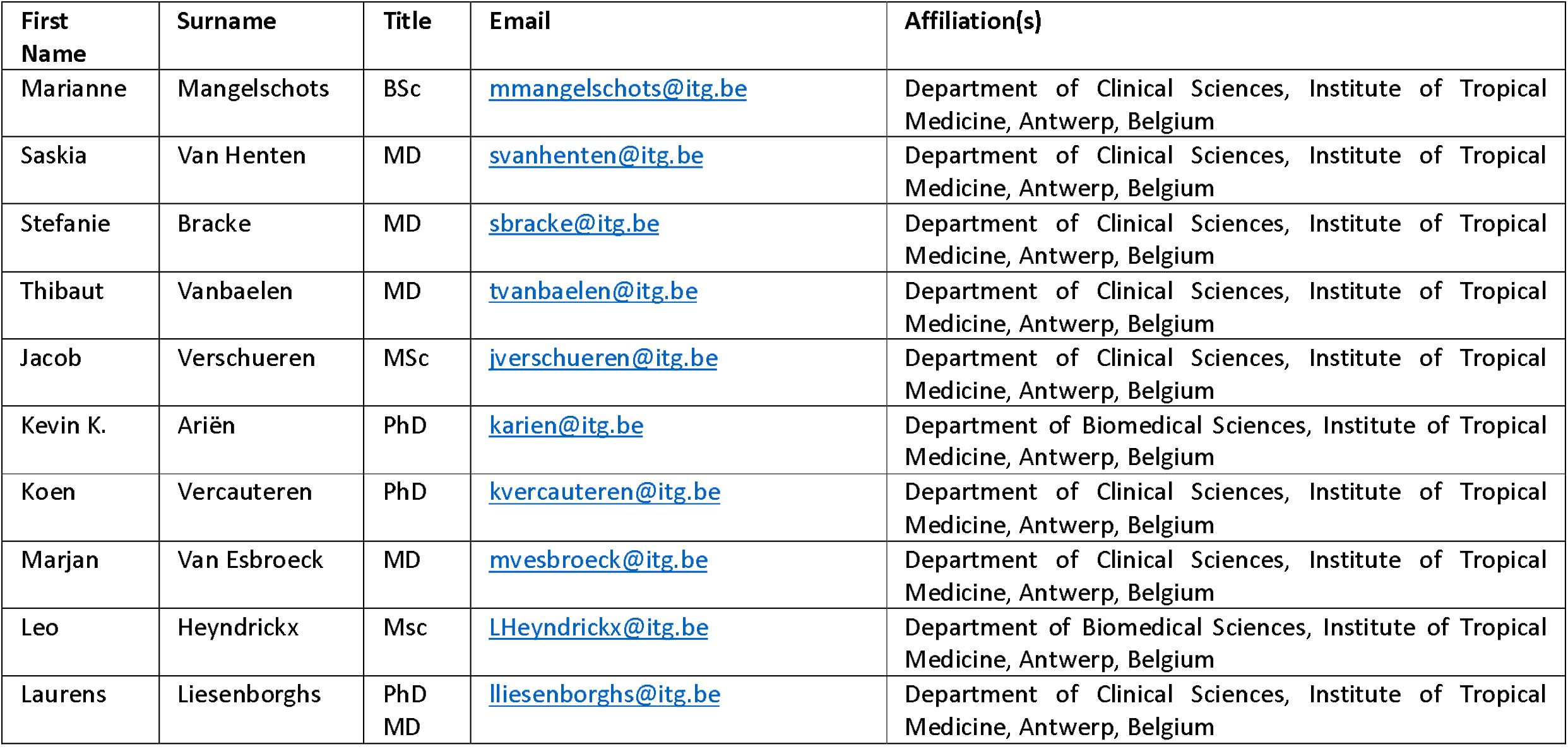

