## Supplementary figures and images for "Pre- and asymptomatic viral shedding in high-risk contacts of monkeypox cases: a prospective cohort study"

### Supplementary Figure 1

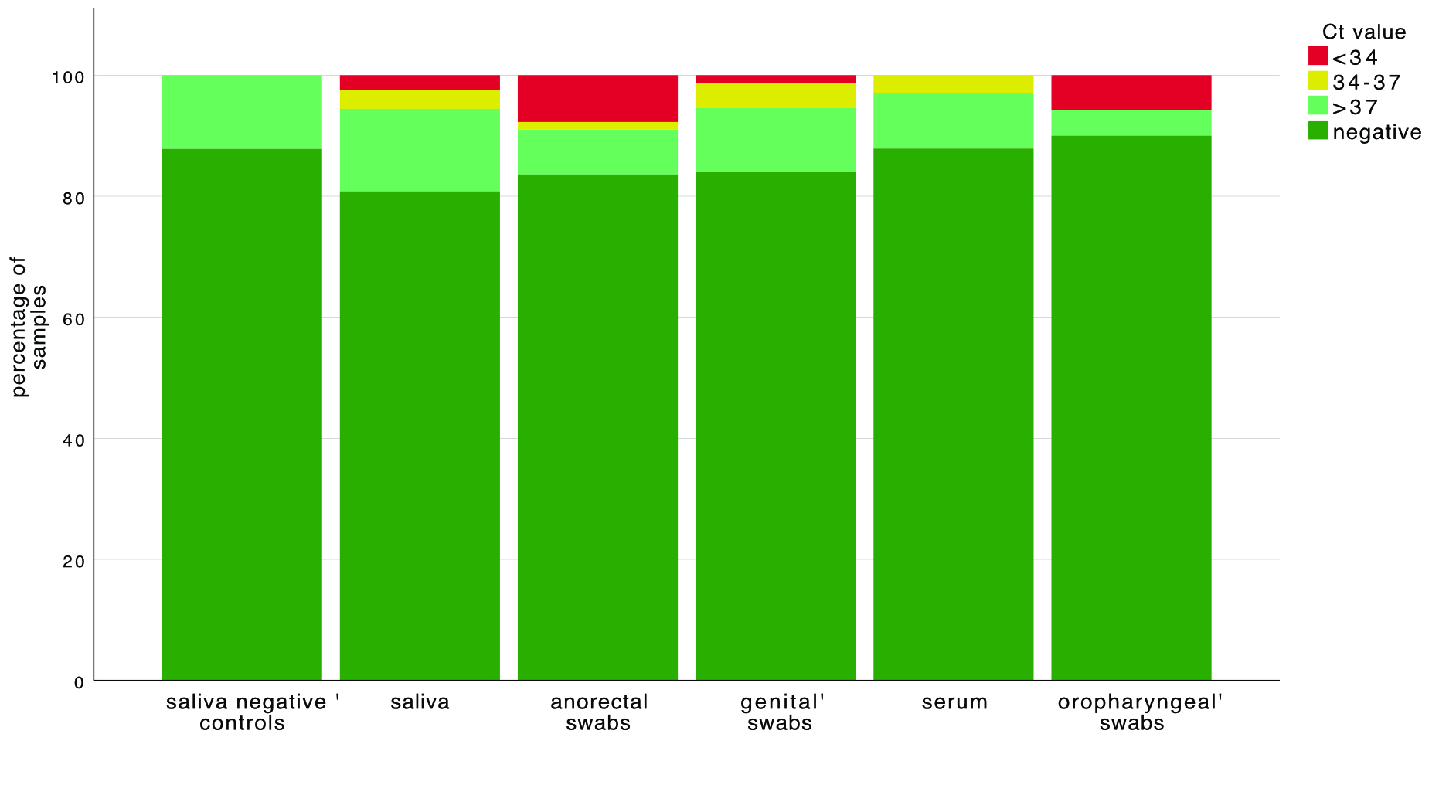

### Supplementary Figure 2

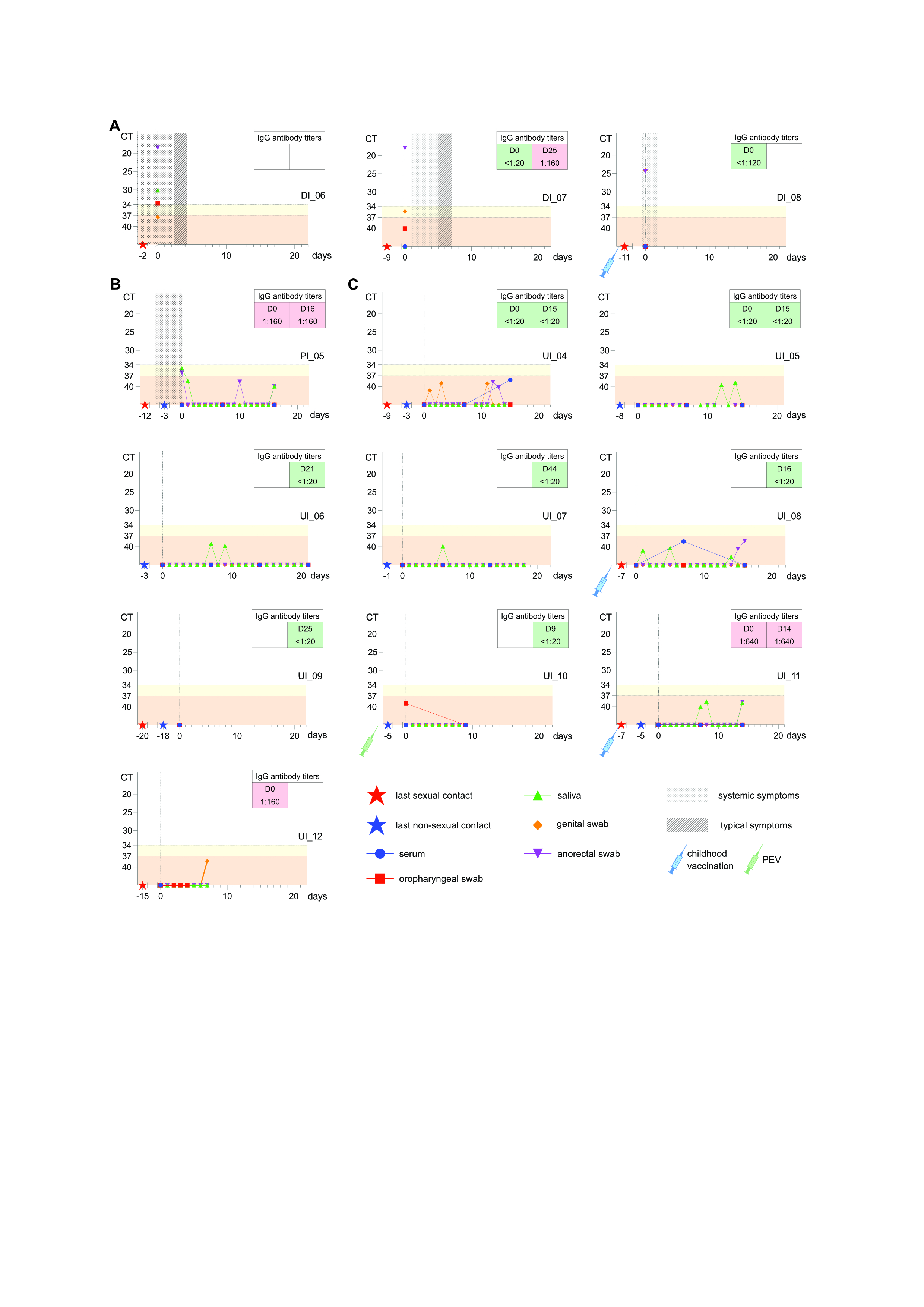
